# Safety and immunogenicity of anti-SARS CoV-2 conjugate vaccine SOBERANA 02 in a two-dose or three-dose heterologous scheme in adults: Phase IIb Clinical Trial

**DOI:** 10.1101/2022.01.01.21268271

**Authors:** María Eugenia Toledo-Romani, Mayra García-Carmenate, Leslyhana Verdecia-Sánchez, Suzel Pérez-Rodríguez, Meybis Rodriguez-González, Carmen Valenzuela-Silva, Beatriz Paredes-Moreno, Belinda Sanchez-Ramirez, Raúl González-Mugica, Tays Hernández-Garcia, Ivette Orosa-Vázquez, Marianniz Díaz-Hernández, María Teresa Pérez-Guevara, Juliet Enriquez-Puertas, Enrique Noa-Romero, Ariel Palenzuela-Diaz, Gerardo Baro-Roman, Ivis Mendoza-Hernández, Yaima Muñoz, Yanet Gómez-Maceo, Bertha Leysi Santos-Vega, Sonsire Fernandez-Castillo, Yanet Climent-Ruiz, Laura Rodríguez-Noda, Darielys Santana-Mederos, Yanelda García-Vega, Guang Wu-Chen, Delaram Doroud, Alireza Biglari, Tammy Boggiano-Ayo, Yury Valdés-Balbín, Daniel García-Rivera, Dagmar García-Rivera, Vicente Vérez-Bencomo, SOBERANA Research Group

**Affiliations:** “Pedro Kourí” Tropical Medicine Institute. Av “Novia del Mediodía”, Kv 6 1/2, La Lisa, Habana, 11400, Cuba; “19 de Abril” Polyclinic. Tulipan Street between Panorama y Oeste, Nuevo Vedado, Plaza de la Revolución, Havana 10400, Cuba; Clinic #1. 21 St. and 190, La Lisa. Havana, Cuba; Finlay Vaccine Institute. 21st Ave. N° 19810 between 198 and 200 St, Atabey, Playa, Havana, Cuba; Cybernetics, Mathematics and Physics Institute. 15th St #55, Vedado, Plaza de la Revolución, Havana10400, Cuba; Centre of Molecular Immunology. 15th Ave. and 216 St, Siboney, Playa, Havana, Cuba; National Civil Defense Research Laboratory. San José de las Lajas, Mayabeque, Cuba; Centre for Immunoassays. 134 St and 25, Cubanacán, Playa, Havana, 11600 Cuba; National Clinical Trials Coordinating Center. 5th Ave and 62, Miramar, Playa, Havana, Cuba; Chengdu Olisynn Biotech. Co. Ltd., and State Key Laboratory of Biotherapy and Cancer Center, West China Hospital, Sichuan University, Chengdu 610041, People’s Republic of China; Pasteur Institute of Iran. No. 69, Pasteur Ave., Tehran 1316943551, Islamic Republic of Iran; Laboratory of Synthetic and Biomolecular Chemistry, Faculty of Chemistry, University of Havana, Havana 10400, Cuba

**Keywords:** conjugate vaccine, heterologous schedule, COVID-19, SARS CoV-2, recombinant RBD, phase IIb clinical trial

## Abstract

**Background:** We report results of immunogenicity, safety and reactogenicity of SOBERANA 02 in a two-dose or three-dose heterologous scheme in adults in a phase IIb clinical trial.

**Method:** This phase IIb trial was designed as parallel, multicentre, adaptive, double blind, randomized and placebo-controlled. Subjects (N=810) aged 19-80 years were randomized to receive two doses of the recombinant SARS CoV-2 receptor binding domain (RBD) conjugated to tetanus toxoid (SOBERANA 02) and a third dose of dimeric RBD (SOBERANA Plus) 28 days apart; two production batches of active ingredient of SOBERANA 02 were evaluated. Primary outcome was the percentage of seroconverted subjects with ≥4-fold the anti-RBD IgG concentration. Secondary outcomes were safety, reactogenicity and neutralizing antibodies.

**Results:** Seroconversion rate in vaccinees was respectively 76.3 and 96.8% after two or three doses, compared with 7.3% in placebo group. Anti-RBD IgG increased significantly after first and second dose of SOBERANA 02 respect to placebo group; and the third dose with SOBERANA Plus boosts the response compared to the second dose. Neutralizing IgG antibodies were detected against D614G and VOCs α, β and δ. Specific and functional antibodies were detected at least until 7-8 months after the third dose. The frequency of serious adverse events (AEs) associated with vaccination was very low (0.1%); with only one serious AE consistent with vaccination. Local pain was the most frequent AE.

**Conclusions:** Two doses of SOBERANA 02 were well tolerated, safe an immunogenic in adults aged 19-80 years old. The heterologous combination with a third dose of SOBERANA Plus increased neutralizing antibodies, detectable 7-8 months after finishing the vaccination schedule.

**Trial registry:** https://rpcec.sld.cu/trials/RPCEC00000347

## INTRODUCTION

COVID-19 disease has led to an unprecedented effort in vaccine development and several vaccines based on different platforms have received emergency use authorization [1]. Despite the outstanding progress, equal access to vaccines continues being a major challenge [2].

SOBERANA 02 is an anti-SARS-CoV-2 vaccine candidate having as immunogen the recombinant RBD protein conjugated to tetanus toxoid [3,4]. Phase I study evaluated safety, reactogenicity and immunogenicity of SOBERANA 02 at two doses (15 and 25 µg) in adults 19-59 years-old and compared SOBERANA 02 in a three-dose (homologous) schedule vs. an heterologous schedule (two doses of SOBERANA 02 and a third dose of SOBERANA Plus, a vaccine candidate whose active pharmaceutical ingredient is the RBD dimer) [5]. After an interim analysis, the higher dose (SOBERANA 02, 25 µg) was selected for phase II study [6] designed in two stages (IIa and IIb); IIa was an open trial evaluating the homologous and heterologous schedules in adults 19-80 years-old [7]. A pooled analysis of both phases I and IIa concluded that the heterologous scheme was safe, well tolerated and elicited the highest immune response, reinforcing memory cells [6].

Here, we report immunogenicity, safety and reactogenicity of two doses of SOBERANA 02 and the heterologous scheme with a third dose with SOBERANA Plus in a randomized, double blind, placebo controlled phase IIb clinical trial.

## MATERIALS AND METHODS

### Participants and study design

Phase IIb was designed as a multicenter, adaptive, parallel, double blind, randomized, placebo controlled trial for evaluating the immunogenicity, safety and reactogenicity of two doses of SOBERANA 02 and the heterologous scheme with a third dose with SOBERANA Plus. Healthy adults aged 19-80 years, of both sexes were recruited through public advertisement at community or professional environment close to the clinical site. Detailed information about all eligibility criteria are summarized in the Supplementary Material Appendix Section A.

Participants were randomly assigned at a 4:4:1 ratio to receive one of the two API batches of SOBERANA 02 or placebo (810 subjects; API 1: 354, API 2: 354 and 102 in the placebo group). Randomization was stratified in four 10-years age subgroups (from 19-29, 30-39, 40-49, 50-59 years), and one 21-years age subgroup (60-80 years).

The trial was conducted at two clinical sites: Clinic #1 at “La Lisa” Municipality and Polyclinic “19 de Abril” at “Plaza de la Revolución” Municipality, Havana, Cuba. (Cuban Public Registry of Clinical Trials, included in WHO International Clinical Registry Trials Platform: *https://rpcec.sld.cu/trials/RPCEC00000347* [7].)

### Products under evaluation

SOBERANA 02 (FINLAY-FR-2) and SOBERANA Plus (FINLAY-FR-1A) are vaccine candidates based on the recombinant receptor binding domain (RBD) of SARS-CoV-2 virus produced in CHO cells. The RBD sequence selected, Arg319-Phe541-(His)_6_, includes free Cys538. This residue provides a suitable conjugation site to tetanus toxoid (in SOBERANA 02) [4]; it also allows dimerization through a disulphide bridge linking two RBD molecules (in SOBERANA Plus) [3]. Vaccines and placebo were produced under Good Manufacturing Practice at Finlay Vaccine Institute and the Center of Molecular Immunology in Havana, Cuba. SOBERANA 02 active pharmaceutical ingredient (API) batches resulted in three batches of the final product: EC-CVRBDC-2003 and EC-CVRBDC-2004 (containing API 1), and EC-CVRBDC-2005 (containing API 2); SOBERANA Plus batches were EC-CVRBDd-2008 and EC-CVRBDd-2101; placebo batch was: E1001PS02. Vaccines composition has been described previously [6]; the placebo contained the excipients of SOBERANA 02. Vaccine and placebo formulations were visually undistinguishable.

### Ethical considerations

The phase II clinical trial protocol was reviewed and approved by an *ad hoc* centralized Research Ethics Committee from the Medical Sciences University, Faculty of Medicine “Manuel Fajardo”, Havana, designed by the Health Innovation Committee from the Cuban Ministry of Health (MINSAP). The Cuban National Regulatory Agency (Center for State Control of Medicines and Medical Devices, CECMED) approved the trial and the procedures (CECMED, Authorization date: 17^th^ December 2020, reference number: 05.019.20BA).

The National Clinical Trials Coordinating Center (CENCEC) was responsible for monitoring the trial in terms of adherence to the protocol and Good Clinical Practice as well as data accuracy. An Independent Data Monitoring Committee (conformed by six external and independent members specialized on clinical practice, epidemiology and statistic) supervised the study.

The trial was conducted following the Declaration of Helsinki, Good Clinical Practice and the rules of the Cuban National Immunization Program. During participants recruitment, the investigators provided the potential participants all relevant information (both orally and written) about the vaccine candidates, and the potential risks and benefits of the trial. All questions and doubts were clarified before enrollment. The decision to participate in the study was voluntary and not remunerated. Written informed consent was obtained from all participants.

### Procedures

Participants received intramuscular injections in the deltoid region, 28 days apart. They were closely followed for one hour after each injection for safety evaluation. Medical visits were planned at 24, 48 and 72 hours, 14 and 28 days after each dose. Any adverse event was self-registered by the participants on a diary card and recorder during medical visits.

Serum samples were collected on days 0 (baseline) and 56 from all subjects; on days 14 and 70, blood samples were taken from 50% of the participants while samples from the other 50% were collected on days 42 and 84. To evaluate the duration of the humoral response, seven to eight months after completing the vaccination schedule, and after signing a new informed consent, another serum sample was obtained from a subset of vaccinated participants.

### Outcomes

The primary outcomes were percentage of subjects with seroconversion ≥4-fold the anti-RBD IgG pre-vaccination level. Secondary outcomes included: 1) Serious Adverse Events (AEs) measured daily for 28 days after each dose; 2) Solicited Local and Systemic AEs for 7 days after each dose; 3) Unsolicited AEs measured daily for 28 days after each dose; 4) Conventional neutralizing antibody titers (cVNT_50_) of a subset of samples from seroconverted subjects and 5) Inhibition of RBD-hACE2 interaction expressed as % and molecular inhibitory titer (mVNT_50_). Outcomes are detailed in Supplementary Material, Appendix A-2).

### Immune response assessment

All immunological evaluations were performed by external laboratories on blind samples. Anti-RBD IgG response and molecular- and conventional-virus neutralization titers were evaluated as described [6] (See Supplementary Materials Appendix C).

Anti-RBD IgG concentration, inhibition of RBD-hACE2 interaction and mVNT_50_ were determined on days 0, 14, 42, 56, 70, 84. Conventional neutralizing antibody titers (cVNT_50_) against D614G variant was evaluated in a subset of samples randomly selected from the individuals with seroconversion on days 0, 56, 70 and 84. cVNT_50_ against VOC was also determined in a subset of samples with cVNT_50_ vs. D614G >20. Molecular neutralization assay (% Inhibition RBD:hACE2) was determined at T0 only if the sample has pre-vaccination IgG concentration over 7.8 AU/ml (4-fold the limit of quantification in ELISA assay, 1.95 AU/ml). Anti-RBD IgG concentration, and mVNT_50_ were also determined after 7-8 months of the last dose.

The humoral immune response was compared with that of a Cuban Convalescent Serum Panel (CCSP) made with serum from 68 COVID-19 convalescent patients and characterized with the same techniques used in clinical trials [6,8].

### Safety evaluation

Solicited local and systemic AEs were measured daily from days 0 to 7 following each immunization. Other AEs were self-recorded until completion of the 28 days follow-up period. The severity of solicited AEs was graded according to Brighton Collaboration definition and the Common Terminology Criteria for Adverse Events version 5·0. All AEs were reviewed for causality, and classified according to WHO [9].

### Statistical analysis

Calculation of sample size was done before starting phase II study (this included phases IIa and IIb) considering a two-sided 95% confidence interval for the difference between two proportions with a width equal to 0.16, to estimate a difference between each API batch and placebo group of around 50%, with a lower bound of the confidence interval greater than 30% and a dropout of 15%. This resulted in a phase II sample size of 910 subjects randomized 4:4:1 in three groups (vaccine API 1, vaccine API 2 and placebo) (404:404:102), and allowing a loss of up to 138 subjects. Stage IIb excluded the 100 participants in phase IIa, giving a sample size of 810 subjects. The evaluation of the study hypothesis remaining valid after excluding stage IIa participants.

Safety and reactogenicity endpoints are described as frequencies (%). Quantitative demographic characteristics are reported as mean, standard deviation (SD), median, interquartile range, and range. We calculated seroconversion rate for anti-RBD IgG antibodies (≥4-fold increase in antibody concentration over baseline) for each subject. Anti-RBD IgG concentration and % of inhibition of RBD-hACE2 interaction were expressed as median and interquartile range; molecular virus neutralization titer (mVNT_50_) and conventional virus neutralization titer (cVNT_50_) were expressed as geometric mean (GMT) and 95% confidence intervals (CI). Spearman’s rank correlation was used to assess relationships among techniques used to evaluate the immune response. The Student t-Test or the Wilcoxon Signed-Rank Test were used for before-after statistical comparison. Statistical analyses were done using SPSS version 25.0; EPIDAT version 12.0 and Prism GraphPad version 6·0. An alpha signification level of 0.05 was used.

## RESULTS

### Flow chart and demographics

From mid-January to end-February 2021, 948 individuals were recruited for phase IIb trial, 138 were excluded and 810 included (Figure 1). Eligible participants were randomly assigned for receiving vaccine (two doses of SOBERANA 02 and one dose of SOBERANA Plus) or placebo, at 28 days intervals.

**Figure 1.**
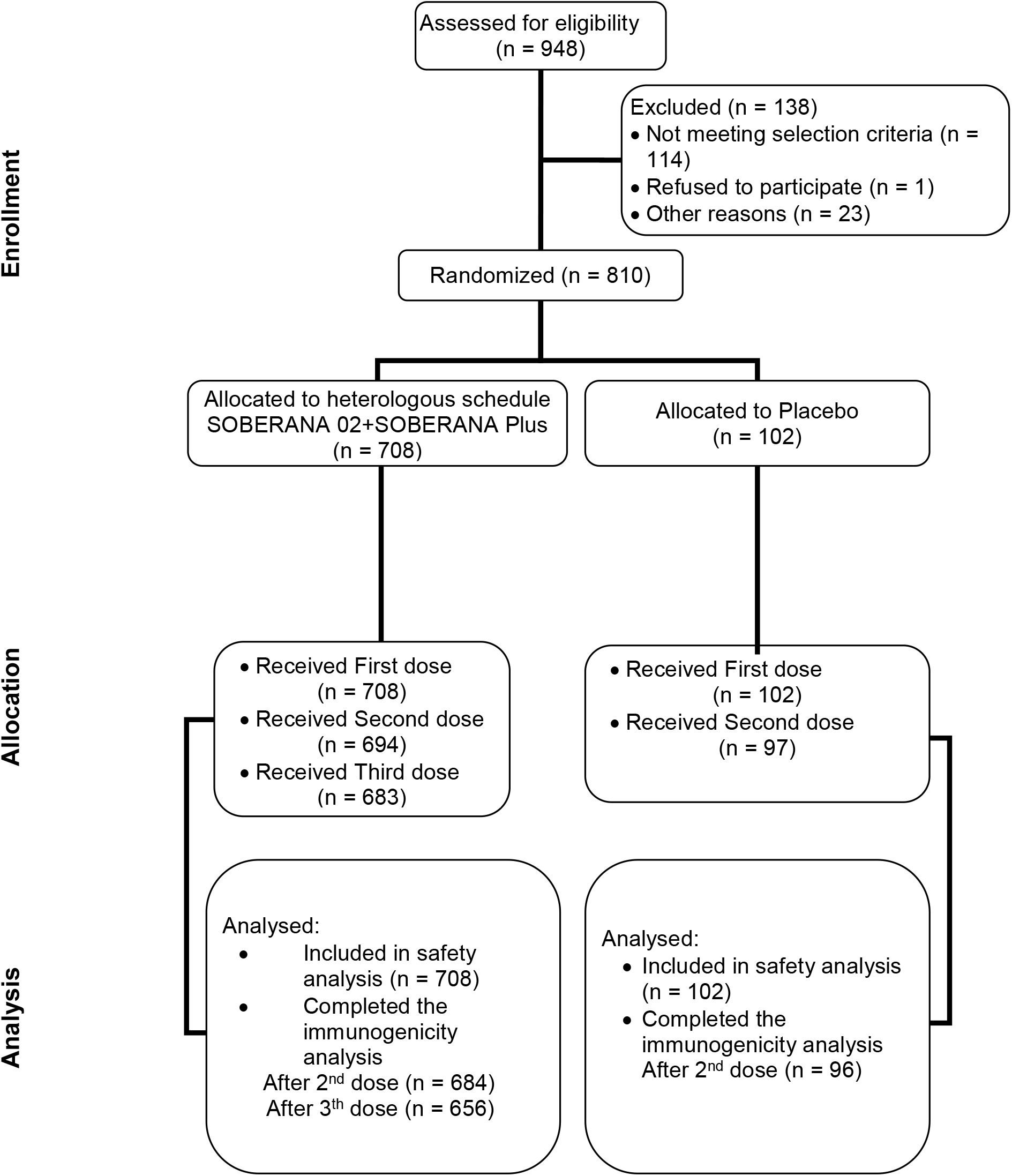
Flow chart of clinical trial phase IIb

The demographics characteristics are summarized in Table 1. The mean age of participants was 47 years (SD 15.8) predominating white race (73.2%).

**Table 1.** Baseline demographic characteristics of subjects included in the clinical trial

|  |  | Groups randomized according to vaccine candidate or placebo |  |  |
| --- | --- | --- | --- | --- |
|  |  | Vaccine | Placebo | Total |
| N |  | 708 | 102 | 810 |
| **Sex** |  |  |  |  |
|  | Female | 359 (50.7%) | 50 (49.0%) | 409 (50.5%) |
|  | Male | 349 (49.3%) | 52 (51.0%) | 401 (49.5%) |
| **Ethnicity** |  |  |  |  |
|  | White | 513 (72.5%) | 80 (78.4%) | 593 (73.2%) |
|  | Black | 84 (11.9%) | 9 (8.8%) | 93 (11.5%) |
|  | Mixed-race | 111 (15.7%) | 13 (12.7%) | 124 (15.3%) |
| **Age (years)** |  |  |  |  |
|  | Mean (SD) | 47.0 ± 15.8 | 47.1 ± 16.0 | 47.0 ± 15.8 |
|  | Median (IQR) | 48.0 ± 26.0 | 48.0 ± 25.0 | 48.0 ± 26.0 |
|  | Range | 19-80 | 19-80 | 19-80 |
|  | 19-59 | 544 (76.8%) | 78 (76.5%) | 622 (76.8%) |
|  | 60-80 | 164 (23.2%) | 24 (23.5%) | 188 (23.2%) |
| **Weight (kg)** |  |  |  |  |
|  | Mean (SD) | 74.2 ± 14.9 | 73.9 ± 12.6 | 74.2 ± 14.6 |
|  | Median (IQR) | 72.0 ± 21.5 | 75.0 ± 18.0 | 73.0 ± 21.0 |
|  | Range | 42-120 | 40-110 | 40-120 |
| **Height (cm)** |  |  |  |  |
|  | Mean (SD) | 167.4 ± 10.0 | 167.0 ± 9.2 | 167.3 ± 9.9 |
|  | Median (IQR) | 167.0 ± 14.0 | 165.5 ± 14.0 | 167.0 ± 14.0 |
|  | Range | 136-200 | 147-186 | 136-200 |
| **BMI (kg/m<sup>2</sup>)** |  |  |  |  |
|  | Mean (SD) | 26.4 ± 4.0 | 26.5 ± 3.8 | 26.4 ± 4.0 |
|  | Median (IQR) | 26.1 ± 5.7 | 26.2 ± 5.4 | 26.1 ± 5.7 |
|  | Range | 18.1-41.0 | 17.3-34.7 | 17.3-41.0 |

Footnote: Vaccine = Heterologous scheme (SOBERANA 02 two doses +SOBERANA Plus). Vaccine data corresponds to results from participants vaccinated with both API batches. Results for individual batches are presented below. Data are n (%) unless otherwise specified. Mean (SD) =Mean ± Standard Deviation. Median (IQR) =Median ± Interquartile Range. BMI=Body mass index.

### Immune response assessment

On day 14, the proportion of subjects with ≥ 4-fold increase in anti-RBD IgG concentration was significantly different (p<0.005) in vaccine (20%) and placebo (3.8%) groups. These values increased to 76.3 % (median 26.5 AU/ml) after second dose (sample on T56) and 96.8% (median 122.2 AU/ml) after third dose (samples collected either on T70 or T84) while values for placebo were 7.3% on T56. This represents a 4.6-fold increase in anti-RBD IgG concentration (p<0.0005) after the third dose compared to the second and a 2.4-fold increase compared to the Cuban Convalescent Serum Panel (CCSP). (Figure 2A, and Supplementary materials Appendix B Table 1).

**Figure 2.**
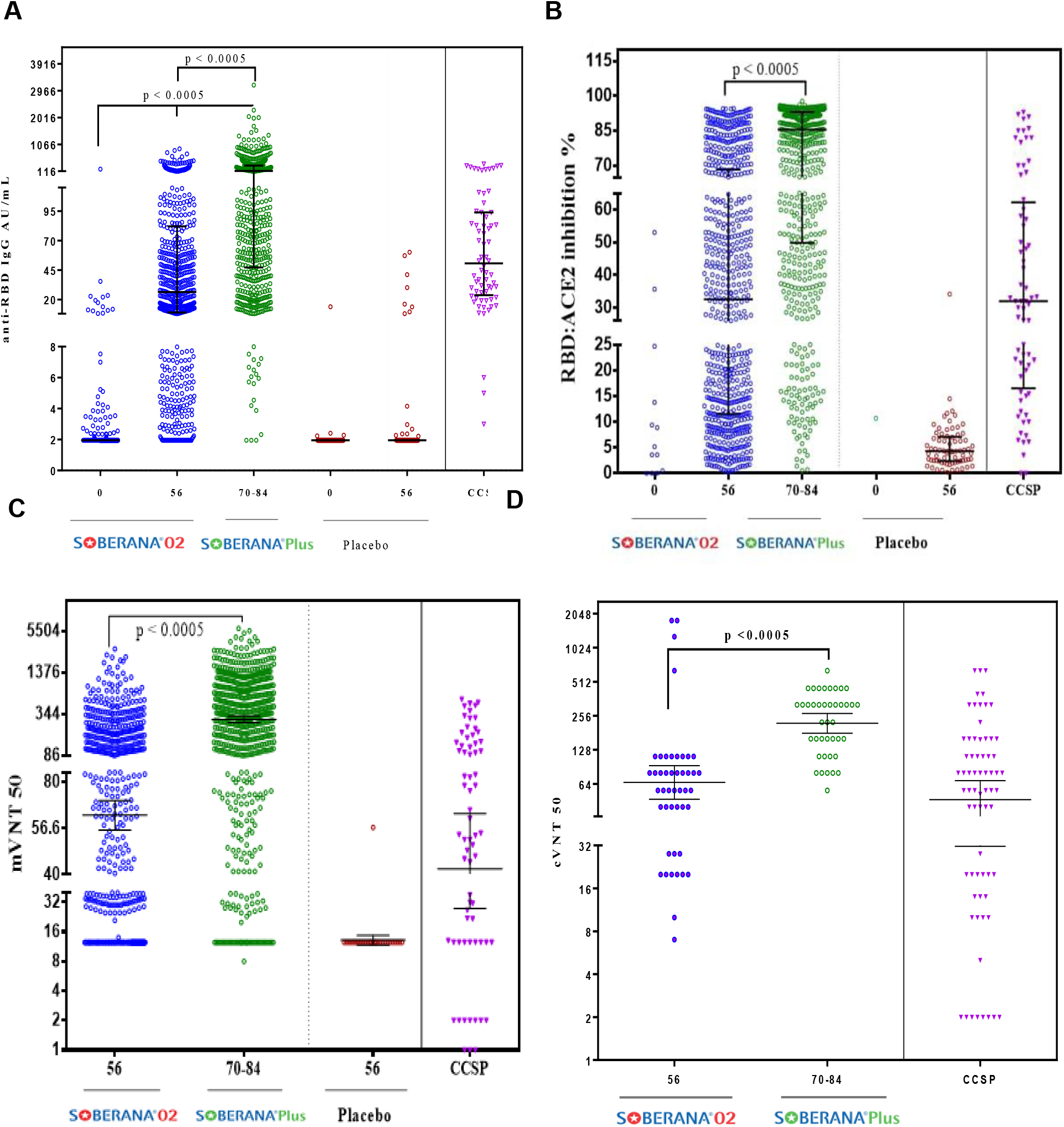
Immunogenicity after vaccination in subjects aged 19-80 years. A) Kinetics of anti-RBD IgG concentration expressed in arbitrary units/mL (median, 25^th^-75^th^ percentile). B) % inhibition of RBD:hACE2 interaction at 1/100 serum dilution (median, 25th-75th percentile). C) Molecular virus neutralization titer mVNT_50_, highest serum dilution inhibiting 50% of RBD:hACE2 interaction (GMT, 95% CI). D) Conventional live-virus neutralization titre cVNT_50_ (GMT, CI 95%) against SARS-CoV-2 D614G variant. Footnote: Blue dots: response after two doses (on T0, T28) of SOBERANA 02. Green dots: response at T70 or T84 after receiving the third dose on T56, this one of SOBERANA Plus. Brown dots: subjects receiving placebo. CCSP (purple): Cuban Convalescent Serum Panel. p value: statistic differences (on T70 orT84) compared to T56 or T0.

The molecular inhibition of RBD:hACE2 interaction (expressed as % inhibition and molecular virus neutralization titer, mVNT_50_) also increased. The inhibition median after two doses of SOBERANA 02 was 28.4% (25^th^-75^th^ percentile 10.8; 67.0), similar to the value for CCSP, 32% (25^th^-75^th^ percentile 26.6; 62.2). After the third dose this value increased to 85.5% (25^th^-75^th^ percentile 49.4; 93.1); the GMT for mVNT_50_ was 289.0 (95% CI: 258.4; 323.4) which represented a 4.6-fold increase compared to the value after the second dose (p<0.0005) and a 6.9-fold increase compared to CCSP value. (Figure 2B-C, Supplementary materials Appendix B Table 1).

The conventional virus neutralization titre (cVNT_50_) was evaluated in a subset of samples randomly selected from participants with seroconversion after second (on T56) and third doses (on days T70 or T84). After two doses, the cVNT_50_ GMT was 65.9 (95% CI 46.9; 92.7), comparable to the CCSP value (GMT 41.8; 95% CI 27.7; 63.2). After the third heterologous dose a remarkable, statistically significant increase (p<0.0005) was observed, attaining a GMT of 219.6 (95% CI 179.5; 268.8); this is 3.3-fold the value after the second dose and 4.7-fold the value of CCSP (Figure 2D, Supplementary materials Appendix B Table 1).

Neutralization against SARS-CoV-2 variants was also analyzed in sera from 18 subjects that completed the vaccination schedule. As seen in Figure 3, cVNT_50_ GMT was 364 (95% CI 304.7; 434.8) against D614G variant, whereas cVNT_50_ GMT of 339 (95% CI 277.3; 414.4), 156.7 (95% CI 122.4; 200.6) and 51.4 (95% CI 31.23; 84.49) were obtained against alpha, delta and beta variants, respectively. Compared to D614G, no differences were detected in neutralizing titres against alpha variant: however, there was a reduction of 2.32-fold and 7-fold against delta and beta variants, respectively. All analysed sera conserved neutralizing activity against alpha and delta variants, and a 94% of sera conserved activity against beta variant.

**Figure 3.**
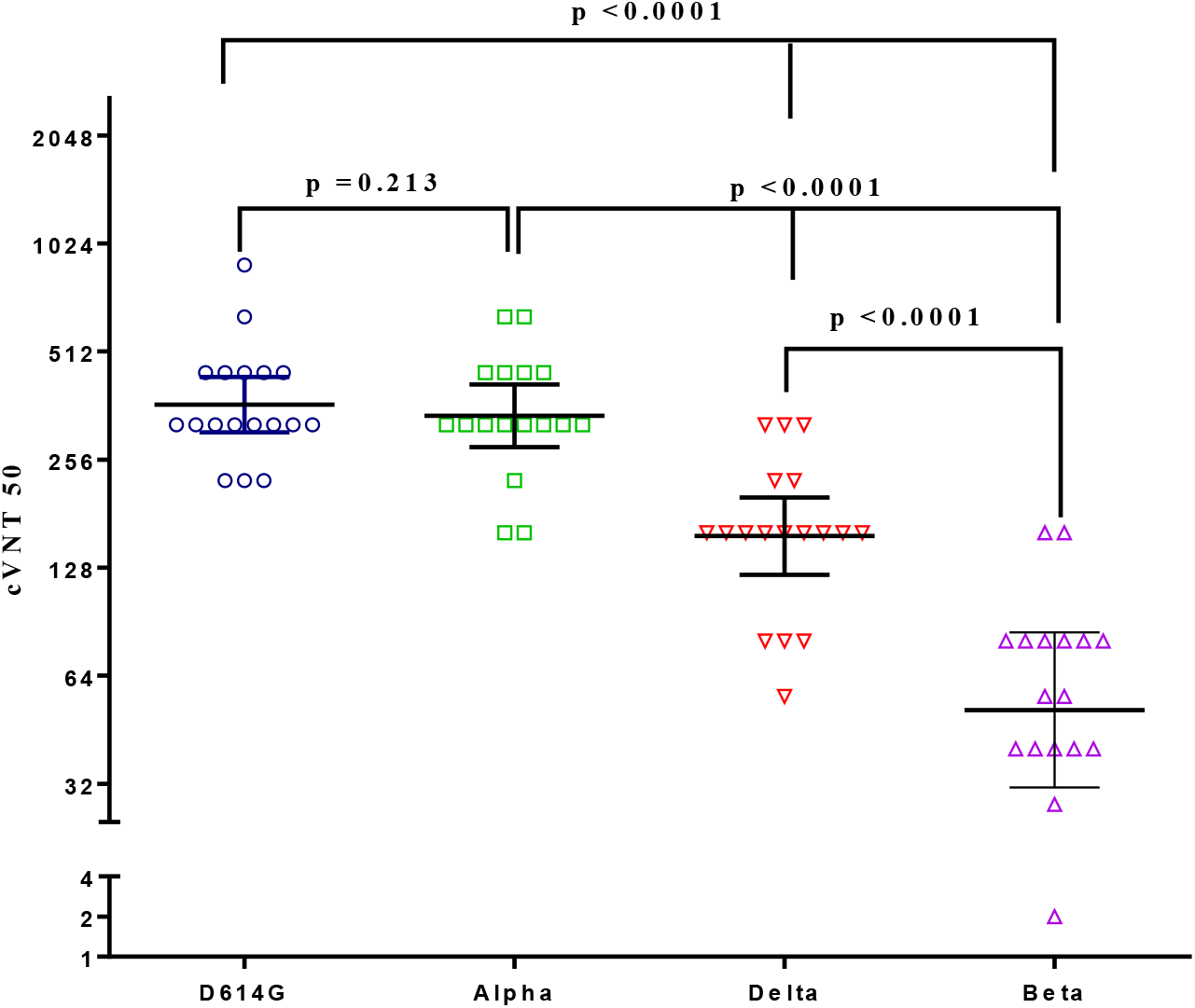
Live-virus neutralization titer against SARS-CoV-2 variants. Sera from 18 subjects vaccinated with complete schedule (two doses SOBERANA 02 + one dose SOBERANA Plus, 28 days apart) were evaluated (cVNT_50_: GMT, 95% CI) against variants B.1.1.7 alpha, B.1.617.2 delta, and B.1.351 beta, and compared with D614G variant. p values represent the statistic differences as indicated, using paired Student t test with log-transformed variables.

In females, in participants 19-59 years and in individuals without high risk comorbidities the analysis of immunological variables by participants’ subgroups indicated significant increase (p<0.00005) in all variables except for conventional virus neutralization titer (cVNT_50_) (Supplementary materials Appendix B, Table 2). Compared to placebo, in the vaccine group there was a significant increase in the immune response for all subgroups (data no shown).

**Table 2.** Main characteristics of adverse events following vaccination

|  | **Groups** |  | **Total** |
| --- | --- | --- | --- |
|  | **Vaccine** | **Placebo** |  |
| **N** | 708 | 102 | 810 |
| **Subjects with some AE** | 336 (47.5%) | 24 (23.5%) | 360 (44.4%) |
| **Subjects with some VAAE** | 311 (43.9%) | 16 (15.7%) | 327 (40.4%) |
| **Subjects with some Serious AE** | 4 (0.5%) | 1 (1.0%) | 5 (0.6%) |
| **Subjects with some Serious VAAE** | 1 (0.1%) | -- | 1 (0.1%) |
| **Subjects with some Severe AE (no VAAE)** | 1 (0.1%) | -- | 1 (0.1%) |
| **Total Adverse Events** | 899 | 48 | 947 |
| **Mild AE** | 831 (92.4%) | 4 (95.8%) | 877 (92.6%) |
| **Moderate AE** | 66 (7.3%) | 2 (2.2%) | 68 (7.2%) |
| **Severe AE** | 2 (0.2%) | -- | 2 (0.2%) |
| **Serious AE** | 7 (0.8%) | 1 (2.1%) | 8 (0.8%) |
| **Local AE** | 583 (64.8%) | 12 (25.0%) | 595 (62.8%) |
| **Systemic AE** | 316 (35.2%) | 36 (75.0%) | 352 (37.2%) |
| **VAAE** | 706 (78.5%) | 26 (54.2%) | 732 (77.3%) |
| **Serious VAAE** | 1 (0.1) | 0 (0.0) | 1 (0.1) |
| **Severe VAAE** | 0 (0.0) | 0 (0.0) | 0 (0.0) |
| **Reported Serious AE (VAAE)** | Multiform erythema | -- |  |

Footnote: Vaccine = Heterologous scheme (SOBERANA 02 two doses +SOBERANA Plus). Vaccine data corresponds to results from participants vaccinated with both API batches. Data are n (%). AE=Adverse Event. VAAE=Vaccine-Associated Adverse Event.

There was a good correlation among all variables (coefficients > 0.8) except for cVNT_50_, this one evaluated in a small number of participants (Supplementary materials Appendix B, Table 3).

**Table 3.** Characterization of Adverse Events

|  | **Vaccine** | **Placebo** |
| --- | --- | --- |
| **N** | 708 | 102 |
| **Overall AE within 28 days** |  |  |
| Subject with AE | 336 (47.5%) | 24 (23.5%) |
| Severe (Grade 3) | 1 (0.1%) | 0 |
| Serious | 4 (0.5%) | 1 (1.0%) |
| **Subjects with solicited AE** |  |  |
| Any | 286 (40.4%) | 13 (12.7%) |
| Severe (Grade 3) | 0 | 0 |
| Serious | 0 | 0 |
| **Subjects with solicited systemic AE** |  |  |
| Any | 32 (4.5%) | 3 (2.9%) |
| General discomfort | 29 (4.1%) | 3 (2.9%) |
| Rash | 1 (0.1%) | 0 |
| Fever | 2 (0.3%) | 1 (1.0%) |
| Mild fever | 1 (0.1%) | 0 |
| **Subjects with solicited local AE** |  |  |
| Any | 294 (38.8%) | 11 (10.8%) |
| Injection-site pain | 251 (35.5%) | 9 (8.8) |
| Erythema | 13 (1.8) | 0 (0.0) |
| Local Warm | 43 (6.1) | 2 (2.0) |
| Induration | 36 (5.1) | 0 (0.0) |
| Swelling | 92 (13.0) | 0 (0.0) |

Footnote: Vaccine = Heterologous scheme (SOBERANA 02 two doses +SOBERANA Plus). Vaccine data corresponds to results from participants vaccinated with both API batches.

Figure 4 shows immunogenicity results in subjects 7-8 months after completing the vaccination schedule. As expected, the specific antibody concentration (median 20.6; 25^th^-75^th^ percentile 6.9; 58.3) decreased significantly (p<0.0001) compared to those after second (24.9; 25^th^-75^th^ percentile 8.2; 85.6) and third dose (121.8; 25^th^-75^th^ percentile 44.5; 343.7) and to CCSP (50.8; 25^th^-75^th^ percentile 23.8; 94.0) (Figure 4A, Supplementary materials Appendix B, Table 4). The proportion of subjects with seroconversion after 7-8 months (73.0%) is similar to that obtained after two doses (74.6%) (Supplementary materials Appendix B, Table 4). Interesting, the mVNT_50_ GMT (149.6; 95% CI: 122.3; 182.9)) was significantly higher than those after second dose and CCSP (Figure 4B, Supplementary materials Appendix B, Table 4).

**Figure 4.**
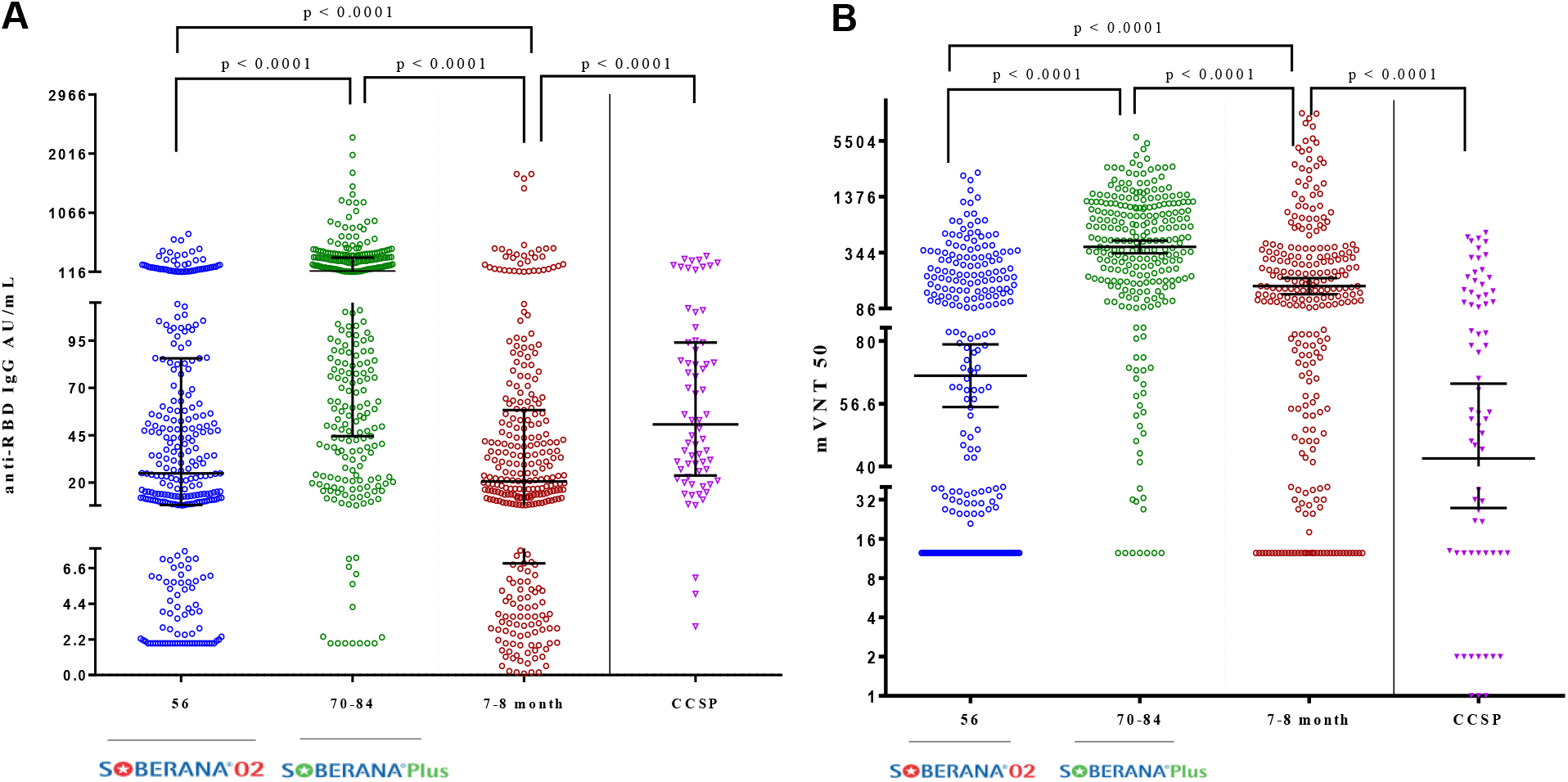
Immunogenicity in vaccinated subjects 7-8 months after completing the immunization schedule (two doses SOBERANA 02 + one dose SOBERANA Plus, 28 days apart). A) Anti-RBD IgG concentration expressed in arbitrary units/mL (median, 25^th^-75^th^ percentile). B) Molecular virus neutralization titer mVNT_50_, highest serum dilution inhibiting 50% of RBD:hACE2 interaction (GMT, 95% CI). Footnote: Blue dots: response after two doses (on T0, T28) of SOBERANA 02. Green dots: response at T70 or T84 after receiving on T56 the third dose, this one SOBERANA Plus (on T). Brown dots: response after 7-8 months. CCSP (purple): Cuban Convalescent Serum Panel. p value: statistic differences.

### Safety analysis

Of the 810 participants, 44.4% presented some AE. In total, 947 AEs of 80 types were reported, 92.6% classified as mild (77.3% consistent with vaccination and 70.7% related to the product under investigation). Eight serious AEs were reported (one —multiform erythema— was consistent with vaccination due to inherent conditions of the subject) (Table 2 and Supplementary materials Appendix B Table 5). The most frequent local solicited AE in both vaccine and placebo groups was pain at injection site (35.5% vs. 8.8%, respectively) followed by swelling (only in vaccine group, 13.0%). General discomfort (4.1% vs. 2.9%, respectively in vaccine and placebo groups) was the most frequent solicited AE at systemic level; other AEs had frequencies < 1% (Table 3). Frequency of unsolicited AEs were 22.5% and 18.6% in vaccine and placebo groups respectively, with headache (5.5%) and hypertension (3.8%) as more recurrent (Supplementary materials Appendix B Tables 7 and 8). One participant in vaccine group died from lung and pancreas neoplasm and pneumonia, classified as serious and severe AEs but no consistent with vaccination. The number of vaccinated subjects reporting AEs decreased after the second and third doses (Supplementary materials Appendix B Figure 1, Table 6).

The likelihood ratio (from Bayes factor) was used as benefit-risk index. We observed a strong evidence for benefit in the vaccine group, (Supplementary materials Appendix B, Figure 2).

### API comparison

The frequency of subjects with AEs as well as their characterization was similar in both subgroups vaccinated with two batches of active pharmaceutical ingredient of SOBERANA 02. The confidence intervals for the immunological tests for both batches have a high intersection, suggesting similarity in the immune response (Supplementary materials Appendix B Tables 9, 10 and 11).

## DISCUSSION

This phase IIb trial confirmed the safety of RBD-tetanus toxoid conjugate (SOBERANA 02) and dimeric RBD (SOBERANA Plus) vaccine candidates in a three-dose heterologous scheme, as already observed in phases I and IIa studies [6]. The proportion of participants with any AE was lower in our study (47.5%) compared to phase I and phase I/II studies for other COVID-19 vaccines produced using several platforms [10,11,12,13,14,15]. In our study, unlike others [11,15,16,17], fever, fatigue or nausea were not reported or were <1%.

The humoral immune response is an important indicator for protection against SARS-CoV-2 [18,19]. The IgG antibody response elicited by vaccination is usually compared to the response induced by natural infection in COVID-19 convalescents [15,17]. Two doses of SOBERANA 02 induced a seroconversion rate of 76.3% and an immune response comparable to the CCSP. The application of a third dose, this one of SOBERANA Plus, increased significantly the number of seroconverted participants to 96.8% as well as the concentration of anti-RBD IgG to 122.2 AU/ml. Previously, we had reported that a single dose of SOBERANA Plus increased several times the neutralizing IgG antibody in COVID-19 convalescents [9]; the third dose of SOBERANA Plus had a similar effect in this heterologous schedule, demonstrating the priming effect of the conjugate vaccine in the two-dose regime, inducing an immunological memory as observed in animal models [4].

The ability of antibodies to inhibit the interaction between recombinant RBD and the human-ACE2 receptor is a proxy for *in vivo* antibody affinity [20]. The virus neutralization titer was comparable to that attained by CCSP, indicating that the antibodies elicited by the immunogens—although representing only a small portion of the viral protein structure—efficiently inhibit its binding to the ACE2 receptor expressed in Vero cells. All these results are consistent with those obtained in pooled analysis of phase I and IIa clinical trials [6].

As seen with other viruses, SARS-CoV-2 have evolved and new variants have been identified, some of them associated with higher transmissibility, mortality and decreased vaccine efficacy [21]. Epidemiology in Havana showed an evolution in variant predominance in 2021, initially D614G, then beta (March-June 2021) and finally delta (July-October 2021) [22]. We found a reduction of cVNT_50_ by 2.32-fold for delta, and 7-fold for beta compared to D614G variant. Similar results have been observed by others: there is a 3-5-fold decrease in neutralizing antibodies against delta compared to alpha in vaccinated subjects [23] and, for the beta variant compared to the original strain, a 7.6-9-fold [24] or 10.3-12.4-fold [25] reduction in neutralization titer was observed in individuals immunized with mRNA vaccines or adenoviral-vectors.

Immune response can be influenced by several factors like age, comorbidities and sex, as noted in other anti-SARS-CoV-2 vaccines [26, 27, 28]. Here, vaccination induced a significant increase in all immunological variables in each subgroup compared to placebo. A significantly higher response, except for cVNT_50_, was observed in female subjects, age group 19-59 and without comorbidities of high risk. In phase IIb the immunological response in elders’ subgroup was lower than that in 19-59 years subgroup, while in phase IIa, there were no differences between both age subgroups, except for mVNT_50_ [6]. This may be related to the smaller number of elderly subjects included in phase IIa (24) compared to the 157 analyzed in phase IIb.

Durability of immune response for anti-COVID-19 vaccines is an issue of outmost importance. In this work, concentration of anti-RBD IgG after 7-8 months decreased 5.9- and 2.44-fold compared to post-third dose and to CCSP values, respectively. However, while molecular neutralization titer also decreased compared to the value after third dose (2.65-fold reduction), the resulting molecular neutralization was significantly higher than after second dose (2.27-fold) or CCSP (3.58-fold), showing the persistence of neutralizing antibodies during that period. Levin et al. observed an 18.3-fold waning of antibody titers in subjects after 6 months of two doses of BNT162b2 vaccine whereas a much lower decrease (4.66-fold) was detected in GMT of neutralizing antibody [29].

A prediction of clinical efficacy has been reported for seven vaccines based on immunogenicity data [30,31]. We used our cumulative data for IgG antibodies and cVNT_50_ from phase I, IIa and IIb (for SOBERANA 02, 25µg; two-dose and heterologous three-dose schedules) to estimate the efficacy, using the same ratio vaccinees vs. CCSP. The efficacy for the two-dose schedule was estimated between 58% and 87% and for the three-dose scheme, between 81% and 93% (Supplementary materials Appendix B, Figure 3). These results have been confirmed during a phase III clinical trial conducted in Havana during March-July 2021, reporting a 71% of efficacy for the two-dose schedule of SOBERANA 02 and 92.4% for the heterologous three-dose schedule [32].

In conclusion, two doses of SOBERANA 02 or SOBERANA 02 + SOBERANA Plus combined in a heterologous schedule were immunogenic, well tolerated and safe in adults aged 19-80 years. The third dose of SOBERANA Plus increased significantly the neutralizing antibody titers. Results obtained here confirmed phase I and IIa results and paved the way for phase III clinical evaluation.

## Data Availability

All data produced in the present study are available upon reasonable request to the authors

## Acknowledgments

We thank Dr. Lila Castellanos for scientific advice. We especially thank all the volunteers who participated in the clinical trial.

