## Supplementary tables and figures for "Safety and immunogenicity of anti-SARS CoV-2 conjugate vaccine SOBERANA 02 in a two-dose or three-dose heterologous scheme in adults: Phase IIb Clinical Trial"

**Supplementary material**

**INDEX**

**Appendix A**
A-1. Selection criteria for phase IIb clinical trial
A-2. Outcomes.

**Appendix B**

**Table 1.** Summary of immunological variables evaluated in participants in the vaccine group, compared to participants in placebo group and the Cuban Convalescent Serum Panel (CCSP)

**Table 2.** Subgroup analysis of immunological variables in vaccine group after three doses
**Table 3.** Correlations between immunological variables after 2 and 3 doses in the vaccine group.
**Table 4.** Summary of immunological variables evaluated in individuals in the vaccine group 7-8 months after receiving the 3rd (heterologous) dose
**Table 5.** Global characterization of adverse events
**Table 6.** Frequency of subjects with adverse event by dose
**Table 7.** Unsolicited adverse events after any injection. Overall safety set
**Table 8.** Unsolicited serious and severe adverse events after any injection. Overall safety set
**Table 9.** Humoral immune response induced in subjects vaccinated with the heterologous schedule by batch of active pharmaceutical ingredient
**Table 10.** Summary of adverse events by batch of active pharmaceutical ingredient
**Table 11.** Frequency of subjects with adverse events by batch of active pharmaceutical ingredient

**Figure 1.** Solicited local and systemic events after each dose.
**Figure 2.** Risk-Benefit Balance.
**Figure 3.** Prediction of clinical efficacy from correlation between antibody responses and efficacy rate

**Appendix C. Immunological Techniques**
C.1 Anti-RBD IgG response
C.2 Molecular virus neutralization tests
C.5 Conventional virus neutralization test

**Appendix A.**

**A1. Selection criteria for phase IIb clinical trial**

Inclusion criteria:

1. Subjects who give written informed consent to participate in the study.

2. Subjects aged 19– 59 years.

3. Women of childbearing potential, using safe contraceptive methods during the study.

4. Physical examination: normal or without clinically significant alterations.

Exclusion criteria:

1. Acute febrile or infectious disease in the 7 days prior to the administration of the vaccine or at the time of its application.

2. Antimicrobial treatment in the 7 days before the administration of the vaccine.

3. Poor weight (BMI $<18.5$) and obesity (BMI $\geq 34.9$).

4. Non-transmissible chronic diseases not controlled according to clinical or laboratory criteria (eg., bronchial asthma, chronic obstructive pulmonary disease, diabetes mellitus, thyroid diseases, ischemic heart disease, arterial hypertension, psychiatric disease, hemolymphopoietic disease).

5. Congenital or acquired immune system disease.

6. History of unresolved neoplastic disease.

7. History of liver or kidney failure.

8. History of substance abuse during the last 30 days or addictive illness to toxic substances, except if the subject is in abstinence, in the case of alcoholics, and smoking.

9. Diminished mental faculties for decision making.

10. History of severe allergic disease (anaphylactic shock, angioneurotic edema, glottis edema, severe urticaria).

11. History of hypersensitivity to thiomersal or to some of the components of the formulation.

12. History of SARS-CoV-2 and COVID-19 who meet any of the following criteria: a) Previous or current history of SARS-CoV-2 infection. b) Be declared in the category of contact or suspect at the time of inclusion. c) Subject with positive test for anti-SARS-CoV-2 antibodies. d) Subject with positive PCR at the time of inclusion.

13. Participation in another clinical trial in the last 3 months.

14. Application of vaccines containing tetanus toxoid in the last 3 months.

15. Application of other vaccines in the last 30 days.

16. Treatment with immunomodulators in the last 30 days, considering steroids (except topical and inhaled), cytostatics, interferon, immunoferon, transfer factor, monoclonal antibody, biomodulina T, any gammaglobulin, Levamisole, Heberferon, thymosin) or other drugs with immunomodulatory action. Persons requiring immunomodulatory treatment during the study.

17. Transfusion of blood or blood products in the last 3 months.

18. Subjects with difficulties in attending the planned follow-up consultations.

19. Splenectomy or splenic dysfunction.

20. Pregnancy, puerperium or breastfeed

21. Tattoos in the deltoid region on both arms.

22. Women with a positive pregnancy test.

**A2- Outcomes.**

*Primary outcome(s):*

1) Anti-RBD IgG antibodies and percentage of subjects with seroconversion ≥4-fold the pre-vaccination level. Data collection: on days 0, 14, 42, 56, 70; 84.

*Secondary outcomes for safety:*

1) Solicited Local and systemic Adverse Events (AE);
   a) Occurrence of the AE (Yes, No)
   b) Duration (hours since the beginning until the end of the event)
   c) Intensity of the AE (mild, moderate, severe)
   d) Severe (serious, not serious)
   e) Result (recovered, recovered with sequelae, persists, death, unknown)
   f) Causality (causal association consistent with vaccination, undetermined, causal association inconsistent with vaccination, not classifiable).
   - Data collection: daily, for 7 days after each dose

2) Unsolicited Adverse Events (AE);
   a) Description of the AE (name of the event)
   b) Duration (hours since the beginning until the end of the event)
   c) Intensity of the AE (mild, moderate, severe)
   d) Severe (serious, not serious)
   e) Result (recovered, recovered with sequelae, persists, death, unknown)

f) Causality (causal association consistent with vaccination, undetermined, causal association inconsistent with vaccination, not classifiable).
- Data collection: daily, for 28 days after each dose

3) Serious Adverse Events (SAEs)
   a) Occurrence of the SAE (Yes, No)
   b) Duration (hours since the beginning until the end of the event)
   c) Description of the event
   d) Result (recovered, recovered with sequelae, persists, death, unknown)
   e) Causality association (causal association consistent with vaccination, inconsistent causal association with vaccination, unclassifiable)
   - Data collection: daily, for 28 days after each dose

*Secondary outcomes for immunogenicity:*

4) $cVNT_{50}$ (conventional live-virus neutralizing titer) Data collection: on days 0, 56, 70 and 84 (sample subset)
5) $mVNT_{50}$ (molecular virus neutralization titer) and % ACE2-RBD inhibition: Data collection: on days 0, 14, 42, 56, 70, 84.

**Appendix B.**

**Table 1.** Summary of immunological variables evaluated in participants in the vaccine group, compared to participants in placebo group and the Cuban Convalescent Serum Panel (CCSP)

| | | Vaccine | | | | | Placebo | | | CCSP |
|---|---|---|---|---|---|---|---|---|---|---|
| | | T0 | Post-1st dose T14 | Post-2nd dose T42 | Post-2nd dose T56 | Post-3rd dose T70-T80 | Post-1st dose T14 | Post-2nd dose T42 | Post-2nd dose T56 | |
| **Anti-RBD IgG seroconversion** | N (%) | - | 72/350 (20.6) | 251/343 (73.2) | 522/684 (76.3) | 635/656 (96.8) | 2/52 (3.8) | 0/46 (0.0) | 7/96 (7.3) | - |
| | 95% CI | - | (16.4; 25.2)* | (68.2; 77.8)* | (72.9; 79.4)* | (95.1; 98.0)* | (0.5; 13.2) | (0.00; 1.3) | (3.0; 14.4) | - |
| **Anti-RBD IgG (AU/mL)** | Median | 1.95 | 2.6 | 28.2 | 26.5 | 122.2 | 1.95 | 1.95 | 1.95 | 50.8 |
| | $25^{th}$-$75^{th}$ | 1.95; 1.95 | 1.95; 6.4* | 7.4; 71.8* | 9.0; 82.2* | 47.2; 314.4* | 1.95; 1.95 | 1.95; 1.95 | 1.95; 1.95 | 23.8; 94.0 |
| **RBD:hACE2 INH%** | Median | 7.0 | 6.3 | 26.1 | 28.4 | 85.5 | 2.7 | 3.7 | 2.8 | 32.0 |
| | $25^{th}$-$75^{th}$ | 0.4; 24.9 | 3.2; 15.0 | 9.4; 63.5* | 10.8; 67.0* | 49.4; 93.1* | 0.5; -- | 1.6; 7.4 | 0.3; 6.3 | 26.6; 62.2 |
| **$mVNT_{50}$** | GMT | - | - | 59.2 | 62.5 | 289.0 | - | - | 13.2 | 41.8 |
| | 95% CI | - | - | 50.5; 69.5* | 55.9; 69.8* | 258.4; 323.4* | - | - | 11.8; 14.7 | 27.7; 63.2 |
| **$cVNT_{50}$** | GMT | 0 | - | - | 65.9 | 219.6 | - | - | - | 46.4 |
| | 95% CI | 0 | - | - | 46.9; 92.7 | 179.5; 268.8 | - | - | - | 31.5; 68.4 |

Footnote: Vaccine = Heterologous scheme: SOBERANA 02 (two doses, 28 days apart) + SOBERANA Plus. AU/mL = anti-RBD IgG concentration expressed in arbitrary units/mL. RBD:hACE2 INH% = RBD:hACE2 inhibition % at a dilution 1/100. $mVNT_{50}$ (molecular virus neutralization test) = serum dilution inhibiting 50% of RBD:hACE2 interaction. $cVNT_{50}$ = conventional live-virus neutralization titre. GMT = Geometric Mean Titre. 25th-75th=25-75 percentile. * $p<0.005$ versus placebo using chi-squared test (anti-RBD IgG seroconversion) Mann-Whitney U (anti-RBD IgG AU/mL, RBD:hACE2 INH%) or Student t test ($mVNT_{50}$, cVNT log-transformed). CCSP = Cuban Convalescent Serum Panel. No $cVNT_{50}$ was performed in samples from placebo as seroconversion was not detected in this group.

**Table 2.** Subgroup analysis of immunological variables in vaccine group after three doses

| | | Conventional viral neutralization ($cVNT_{50}$) | Anti-RBD IgG concentration | Inhibition (%) of RBD:hACE2 interaction | Molecular virus neutralization ($mVNT_{50}$) |
|---|---|---|---|---|---|
| | | GMT (CI 95%) | Median ($25^{th}$-$75^{th}$ Percentile) | Median ($25^{th}$-$75^{th}$ Percentile) | GMT (CI 95%) |
| Sex | F | N=25 | N=334 | N=334 | N=333 |
| | | 198.7 (148.1; 266.5) | 159.4 (60.2; 357.4) | 89.6 (62.0; 93.7) | 369.6 (317.2; 430.7) |
| | M | N=16 | N=322 | N=322 | N=320 |
| | | 256.9 (198.2; 333.2) | 96.5 (37.6; 261.4) | 79.7 (39.9; 91.6) | 223.8 (190.6; 262.8) |
| | Mann-Whitney U | 0.178* | 0.000038 | 8.5e-7 | 0.00001* |
| Age group | 19-59 y/o | N=38 | N=497 | N=497 | N=496 |
| | | 229.4 (185.9; 282.9) | 162.5 (62.1; 359.4) | 88.5 (61.9; 93.5) | 357.3 (316.5; 403.5) |
| | 60-80 y/o | N=3 | N=159 | N=159 | N=157 |
| | | 127.0 (47.0; 343.2) | 53.5 (19.8; 141.1) | 59.7 (29.5; 87.4) | 147.9 (116.5; 187.8) |
| | Mann-Whitney U | 0.125* | 2.5e-14 | 3.4e-12 | 2.0e-11* |
| C | Yes | N=6 | N=225 | N=225 | N=224 |
| | | 189.2 (96.0; 372.8) | 78.6 (24.0; 252.5) | 76.9 (33.0; 92.4) | 202.1 (163.5; 187.8) |
| | No | N=35 | N=431 | N=431 | N=429 |
| | | 225.3 (180.6; 281.2) | 160.1 (61.9; 351.9) | 87.9 (62.0; 93.5) | 348.4 (306.8; 395.8) |
| | Mann-Whitney U | 0.544* | 1.8e-7 | 0.000003 | 0.000018* |

Footnote: C: comorbidities. * Student t test N: number of participants' sera evaluated.

**Table 3.** Correlations between immunological variables after 2 and 3 doses in the vaccine group.

| | | | Anti-RBD IgG (UA/mL) | % Inh RBD:hACE2 | $mVNT_{50}$ |
|---|---|---|---|---|---|
| **After 2 doses** | **Anti-RBD IgG UA/mL** | $r^2$ Spearman | 1.000 | | |
| | | p | . | | |
| | | N | 684 | | |
| | **% Inh RBD:hACE2** | $r^2$ Spearman | 0.918** | 1.000 | |
| | | p | 0.000 | . | |
| | | N | 681 | 681 | |
| | **$mVNT_{50}$** | $r^2$ Spearman | 0.899** | 0.965** | 1.000 |
| | | p | 0.000 | 0.000 | . |
| | | N | 632 | 632 | 632 |
| | **$cVNT_{50}$** | $r^2$ Spearman | 0.598** | 0.629** | 0.632** |
| | | p | 0.000 | 0.000 | 0.000 |
| | | N | 46 | 46 | 46 |
| **After 3 doses** | **Anti-RBD IgG UA/mL** | $r^2$ Spearman | 1.000 | | |
| | | p | . | | |
| | | N | 656 | | |
| | **% Inh RBD:hACE2** | $r^2$ Spearman | 0.900** | 1.000 | |
| | | p | 0.000 | . | |
| | | N | 656 | 656 | |
| | **$mVNT_{50}$** | $r^2$ Spearman | 0.909** | 0.948** | 1.000 |
| | | p | 0.000 | 0.000 | . |
| | | N | 653 | 653 | 653 |
| | **$cVNT_{50}$** | $r^2$ Spearman | 0.787** | 0.618** | 0.729** |
| | | p | 0.000 | 0.000 | 0.000 |
| | | N | 41 | 41 | 41 |

Footnote: ** p<0.01

**Table 4.** Summary of immunological variables evaluated in individuals in the vaccine group 7-8 months after receiving the 3rd (heterologous) dose

| | | Vaccine | | | CCSP |
|---|---|---|---|---|---|
| | | Post-2nd dose T56 | Post-3rd dose T70-T80 | 7-8 months post 3rd dose | |
| **Anti-RBD IgG seroconversion** | N | 307 | 307 | 307 | |
| | N (%) | 229 (74.6) | 291 (94.8) | 224 (73.0) | - |
| | 95% CI | (69.3; 79.4) | (91.7; 97.0)* | (67.6; 77.8) | - |
| **Anti-RBD IgG (AU/mL)** | N | 307 | 307 | 307 | 68 |
| | Median | 24.9 | 121.8 | 20.6 | 50.8 |
| | $25^{th}$-$75^{th}$ | 8.2; 85.6* | 44.5; 343.7* | 6.9; 58.3 | 23.8; 94.0 |
| **$mVNT_{50}$** | N | 261 | 261 | 261 | 68 |
| | GMT | 66.0 | 396.7 | 149.6 | 41.8 |
| | 95% CI | 55.5; 78.5* | 338.7; 464.6* | 122.3; 182.9 | 27.7; 63.2 |

Footnote: Vaccine = Heterologous scheme of SOBERANA 02 + SOBERANA Plus. AU/mL=anti-RBD IgG concentration expressed in arbitrary units/mL. mVNT50 = serum dilution inhibiting 50% of RBD:hACE2 interaction. GMT=Geometric Mean Titre. $25^{th}$-$75^{th}$ = 25-75 percentile. * p<0.005 versus 7-8 months using McNemar test (anti-RBD IgG titer Wilcoxon Signed Ranks test (anti-RBD IgG AU/mL) or Paired Student t test ($mVNT_{50}$, log-transformed). CCSP=Cuban Convalescent Serum Panel.

**Table 5.** Global characterization of adverse events

| | | Vaccine | | Placebo | |
|---|---|---|---|---|---|
| | | Frequency | % | Frequency | % |
| Total adverse events | | 899 | 100.0 | 48 | 100.0 |
| Intensity | Mild (Grade 1) | 831 | 92.4 | 46 | 95.8 |
| | Moderate (Grade 2) | 66 | 7.3 | 2 | 2.2 |
| | Severe (Grade 3) | 2 | 0.2 | 0 | 0.0 |
| | *lung and pancreas neoplasm (1), pneumonia (1)* | | | | |
| Severity | No Serious | 892 | 99.2 | 47 | 97.9 |
| | Serious* | 7 | 0.8 | 1 | 2.1 |
| Causal relationship** | Consistent (A1) | 649 | 72.2 | 21 | 43.8 |
| | Consistent (A2) | 54 | 6.0 | 5 | 10.4 |
| | Consistent (A3) | 3 | 0.3 | 0 | 0.0 |
| | Indeterminate (B1) | 16 | 1.8 | 2 | 4.2 |
| | Indeterminate (B2) | 3 | 0.3 | 0 | 0.0 |
| | Inconsistent | 175 | 19.5 | 20 | 41.7 |
| Result | Recovered | 890 | 99.0 | 48 | 100.0 |
| | Recovered with sequels | 1 | 0.3 | 0 | 0.0 |
| | | *Swelling* | | | |
| | Persist | 8 | 0.9 | 0 | 0.0 |
| | *lung and pancreas neoplasm, diarrhea (2), myalgia, gallstones paint, glossitis, scabies. pneumonia* | | | | |
| Type | Local | 583 | 64.8 | 12 | 25.0 |
| | Systemic | 316 | 35.2 | 36 | 75.0 |
| Solicited | Solicited | 614 | 68.3 | 16 | 33.3 |
| | Unsolicited | 285 | 31.7 | 32 | 66.7 |
| Starting at | ≤ 60 min | 45 | 5.0 | 4 | 8.3 |
| | 60 min-24 hours | 516 | 57.4 | 21 | 43.8 |
| | 24-48 hours | 223 | 24.8 | 12 | 25.0 |
| | 48-72 hours | 61 | 6.8 | 6 | 12.5 |
| | > 72 hours | 54 | 6.0 | 5 | 10.4 |
| Duration (hours) | ≤ 24 hours | 608 | 67.6 | 35 | 72.9 |
| | 24-48 hours | 150 | 16.7 | 4 | 8.3 |
| | 48-72 hours | 74 | 8.2 | 1 | 2.1 |
| | > 72 hours | 53 | 5.9 | 7 | 14.6 |
| | Unspecified | 14 | 1.6 | 1 | 2.1 |

Footnote: Frequency: number of events, %: values referred to total events. * In Vaccine group: Consistent to conditions inherent to the vaccinated *(*multiform erythema); Inconsistent (pulmonary neoplasm, pneumonia, diarrhea, lipothymia, rectal bleeding, atrial fibrillation). In placebo group: Inconsistent (megaloblastic anemia).

** *A1:* Vaccine-related event (according to published literature)*; A2:* Event related to a defect in the quality of the vaccine*; A3:* Event related to a programmatic / technical error*; A4:* Event related to the conditions inherent to the vaccinated subject*; B1:* The temporal relationship is consistent, but the evidence is insufficient to consider the vaccination as cause of the event*; B2:* Classification criteria result in contradiction regarding consistencies and inconsistencies with a causal association with immunization.

**Table 6.** Frequency of subjects with adverse event by dose

| | Any adverse event | |
|---|---|---|
| | **Vaccine Frequency (%)** | **Placebo Frequency (%)** |
| **N** | 758 | 102 |
| **Overall adverse events within 28 days after vaccination** | | |
| Subject with AE | 352 (46.4) | 24 (23.5) |
| 1st dose | 312/758 (41.2) | 21/102 (20.6) |
| 2nd dose | 169/743 (22.7) | 8/97 (8.2) |
| 3rd dose | 64/732 (8.7) | -- |
| **Subjects with solicited adverse events within 7 days after vaccination** | | |
| Any | 343 (45.3) | 22 (21.6) |
| 1st dose | 305/758 (40.2) | 18/102 (17.6) |
| 2nd dose | 164/743 (22.1) | 8/97 (8.2) |
| 3rd dose | 64/732 (8.7) | -- |
| **Subjects with solicited systemic adverse events** | | |
| Any | 35 (4.6) | 3 (2.9) |
| 1st dose | 20/758 (2.6) | 1/102 (1.0) |
| 2nd dose | 12/743 (1.6) | 2/97 (2.1) |
| 3rd dose | 7/732 (1.0) | -- |
| General discomfort | 32 (4.2) | 3 (2.9) |
| 1st dose | 19/758 (2.6) | 1/102 (1.0) |
| 2nd dose | 10/743 (1.3) | 2/97 (2.1) |
| 3rd dose | 7/732 (1.0) | -- |
| Rash | 1 (0.1) | 0 |
| 1st dose | 1/758 (0.1) | 0 |
| 2nd dose | 0 | 0 |
| 3rd dose | 0 | -- |
| Fever | 2 (0.3) | 1 (1.0) |
| 1st dose | 0 | 0 |
| 2nd dose | 1/743 (0.1) | 1/97 (1.0) |
| 3rd dose | 1/732 (0.1) | -- |
| Mild fever | 1 (0.1) | 0 |
| 1st dose | 0 | 0 |
| 2nd dose | 1/743 (0.1) | 0 |
| 3rd dose | 0 | -- |
| **Subjects with solicited local adverse events** | | |
| Any | 294 (38.8) | 11 (10.8) |
| 1st dose | 245/758 (32.3) | 9/102 (8.8) |
| 2nd dose | 150/743 (20.2) | 2/97 (2.1) |
| 3rd dose | 51/732 (7.0) | -- |
| Injection-site pain | 263 (34.7) | 9 (8.8) |
| 1st dose | 223/758 (29.4) | 7/102 (6.9) |
| 2nd dose | 131/743 (17.6) | 2/97 (2.1) |
| 3rd dose | 24/732 (3.3) | -- |
| Erythema | 13 (1.7) | 0 (0.0) |

| | Any adverse event | |
|---|---|---|
| | **Vaccine Frequency (%)** | **Placebo Frequency (%)** |
| **N** | 758 | 102 |
| 1st dose | 7/758 (0.9) | 0 |
| 2nd dose | 2/743 (0.3) | 0 |
| 3rd dose | 4/732 (0.5) | -- |
| Local Warm | 44 (5.8) | 2 (2.1) |
| 1st dose | 24/758 (3.2) | 2 (2.1) |
| 2nd dose | 11/743 (1.5) | 0 |
| 3rd dose | 12/732 (1.6) | -- |
| Induration | 37 (4.9) | 0 (0.0) |
| 1st dose | 22/758 (2.9) | 0 |
| 2nd dose | 13/743 (1.7) | 0 |
| 3rd dose | 6/732 (0.8) | -- |
| Swelling | 93 (12.3) | 0 (0.0) |
| 1st dose | 51/758 (6.7) | 0 |
| 2nd dose | 32/743 (4.3) | 0 |
| 3rd dose | 27/732 (3.7) | -- |

**Table 7.** Unsolicited adverse events after any injection. Overall safety set

| | **Vaccine Frequency (%)** | **Placebo Frequency (%)** |
|---|---|---|
| **N** | 708 | 102 |
| Number of participants reporting unsolicited AEs | 159 (22.5) | 19 (18.6) |
| Vaccine-related | 72 (10.2)* | 7 (6.9)** |
| Hypertension | 27 (3.8) | 3 (2.9) |
| Headache | 39 (5.5) | 2 (2.0) |
| Non Vaccine-related | 114 (16.1)*** | 15 (14.7)**** |
| Hypertension | 7 (1.0) | 1 (1.0) |
| Headache | 18 (2.5) | 1 (1.0) |
| Upper Acute respiratory infection | 14 (2.0) | 3 (2.9) |
| Diarrhea | 17 (2.4) | 0 |
| Myalgia | 18 (2.5) | 1 (1.0) |
| Abdominal pain | 11 (1.6) | 1 (1.0) |

Footnote: * Reported by 1 subject (0.1%, each one): burning, asthenia, nasal secretion, diarrhea, precordial pain, multiform erythema, chills, general discomfort, dizziness, myalgia, migraine, nausea, toothache, paresthesia in upper limb, blush, drowsiness, tachycardia, cough, vertigo, vomiting; local hematoma (3 subjects: 0.4%).

** Reported by 1 subject (1%, each one): general discomfort, palpitations

*** Reported by < 1%: dental abscess, oral thrush, food allergy, anxiety, burning when urinating, asthenia, volume increase in the abdominal region, sinus bradycardia, cellulite in hand and arm, conjunctivitis, nasal secretion, atopic dermatitis, pain by vesicular lithiasis, precordial pain, chest pain, edema in lower limbs, bilateral epicondylitis, scabies, chills, pharyngeal burning, allergic pharingitis, atrial fibrillation, knee forunculus, glositis, stocks with blood, hydroadenitis, acute ingestion, acute lymphangitis in lower member, lipotimia, general discomfort, eye disorder, dizziness, migraine, nausea, lung and pancreas neoplasm, pneumonia, ,tooth pain, osteocondritis, paresthesia, paresthesia in upper limb, paronychia, insect bite pain, polaquyuria, itching in upper limb, generalized itching, local itching, allergic rhinitis, blush, salpingitis, rectal bleeding, urinary sepsis, vagal syndrome, drowsiness, stiff neck, cough, vertigo, vomiting.

****Reporting for 1 subject (1%, each one): submaxilar adenitis, megaloblastic anemia, sinus bradycardia, cellulite in sacral region, conjunctivitis, atopic dermatitis, pharyngeal itching, dizziness, migraine, otalgia palpitations, blush, drowsiness

**** Reporting for 1 subject (1%, each one): Submaxilar adenitis, Megaloblastic anemia, Sinus bradycardia, Cellulite in sacral region, Conjunctivitis, Atopic dermatitis, Pharyngeal itching, Dizziness, Migraine, Otalgia Palpitations, Blush, Drowsiness

**Table 8.** Unsolicited serious and severe adverse events after any injection. Overall safety set

| | **Vaccine** | **Placebo** |
|---|---|---|
| | 708 | 102 |
| Number of participants reporting unsolicited AEs | 159 (22.5) | 19 (18.6) |
| Number of participants reporting unsolicited Serious AEs | 4 (0.6%) | 1 (1.0%) |
| Vaccine-related | 1 (0.1) | 0 |
| Multiform erythema | 1 (0.1) | 0 |
| Non Vaccine-related | 3 (0.4) | 1 (1.0) |
| Atrial fibrillation | 1 (0.1) | 0 |
| Lung and pancreas neoplasm[S1] | 1 (0.1) | 0 |
| Pneumonia[S1] | 1 (0.1) | 0 |
| Lipothymia[S2] | 1 (0.1) | 0 |
| Rectal bleeding [S2] | 1 (0.1) | 0 |
| Diarrhea [S2] | 1 (0.1) | 0 |
| Megaloblastic anemia | 0 | 1 (1.0) |
| Number of participants reporting unsolicited Severe AEs | 1 (0.1) | 0 |
| Non Vaccine-related | 1 (0.1) | 0 |
| Lung and pancreas neoplasm[S1] | 1 (0.1) | 0 |
| Pneumonia[S1] | 1 (0.1) | 0 |

Footnote: [S1]: In the same subject; [S2]: In the same subject

**Table 9.** Humoral immune response induced in subjects vaccinated with the heterologous schedule by batch of active pharmaceutical ingredient

| | | API Batch-1 | | API Batch-2 | |
|---|---|---|---|---|---|
| | | Post-2$^{nd}$ dose T56 | Post-3$^{rd}$ dose T70-T80 | Post-2$^{nd}$ dose T56 | Post-3$^{rd}$ dose T70-T80 |
| **Anti-RBD IgG seroconversion** | N (%) | 265/341 (77.7) | 307/318 (96.5) | 257/343 (74.9) | 328/338 (97.0) |
| | 95% CI | (72.9; 82.0) | (93.9; 98.3) | (70.0; 79.4) | (94.6; 98.6) |
| **Anti-RBD IgG** | Median | 32.7 | 121.9 | 22.5 | 128.4 |
| **(AU/mL)** | 25$^{th}$-75$^{th}$ | 10.1; 98.6 | 45.3; 328.4 | 8.1; 63.5 | 48.9; 311.4 |
| **RBD:hACE2 INH%** | Median | 35.2 | 84.7 | 24.2 | 86.4 |
| | 25$^{th}$-75$^{th}$ | 11.2; 71.6 | 45.1; 92.9 | 10.4; 58.0 | 52.2; 93.2 |
| **mVNT$_{50}$** | GMT | 70.2 | 275.1 | 55.4 | 302.7 |
| | 95% CI | 59.8; 82.4 | 232.5; 325.4 | 47.6; 64.5 | 260.5; 351.8 |
| **cVNT$_{50}$** | GMT | 69.9 | 201.8 | 61.9 | 242.4 |
| | 95% CI | 42.9; 113.7 | 153.4; 265.3 | 36.9; 103.8 | 175.7; 334.2 |

Footnote: API: Active Pharmaceutical Ingredient, AU/mL=anti-RBD IgG concentration expressed in arbitrary units/mL. RBD:hACE2 INH%= RBD:hACE2 inhibition % at a dilution 1/100. mVNT$_{50}$=serum dilution inhibiting 50% of RBD:hACE2 interaction. cVNT$_{50}$=conventional live-virus neutralization titer. GMT=Geometric Mean Titre. 25th-75th=25-75 percentile. CI=confidence interval

**Table 10.** Summary of adverse events by batch of active pharmaceutical ingredient

| | API Batch-1 | API Batch-2 |
|---|---|---|
| **N (%)** | **354 (100.0)** | **354 (100.0)** |
| Subjects with some AE | **164 (46.3)** | **172 (48.6)** |
| Subjects with some VAAE | 157 (44.4) | 154 (43.5) |
| Subjects with some Serious AE | 2 (0.6) | 2 (0.6) |
| Subjects with some Serious VAAE | 1 (0.3) | 0 (0.0) |
| Subjects with some Severe AE (no VAAE) | 1 (0.3) | 0 (0.0) |
| **Total Adverse Events** | **429 (100)** | **470 (100)** |
| VAAE | 339 (79.0) | 367 (78.1) |
| Serious AE | 3 (0.7) | 4 (0.9) |
| Serious VAAE | 1 (0.2) | 0 (0.0) |
| Severe AE(no VAAE) | 2 (0.5) | 0 (0.0) |

Footnote: API: Active Pharmaceutical Ingredient, VAAE: Vaccine-Associated Adverse Event

**Table 11** Frequency of subjects with adverse events by batch of active pharmaceutical ingredient

| | | | API Batch-1 | API Batch-2 |
|---|---|---|---|---|
| **Total (%)** | | | **354 (100.0)** | **354 (100.0)** |
| Subjects with some AE | | | **164 (46.3)** | **172 (48.6)** |
| Solicited | Local | Injection-site pain | 123 (34.7) | 128 (36.2) |
| | | Swelling | 46 (13.0) | 46 (13.0) |
| | | Local warm | 21 (5.9) | 22 (6.2) |
| | | Induration | 16 (4.5) | 20 (5.6) |
| | | Erythema | 6 (1.7) | 7 (2.0) |
| | Systemic | General discomfort | 18 (5.1) | 11 (3.1) |
| | | Fever | 1 (0.3) | 1 (0.3) |
| | | Rash | 1 (0.3) | 0 (0.0) |
| | | Mild fever | 0 (0.0) | 1 (0.3) |
| Unsolicited | Local | Local Hematoma | 3 (0.8) | 0 (0) |
| | | Local Itching | 1 (0.3) | 0 (0) |
| | | Burning | 1 (0.3) | 0 (0) |
| | | Cellulite of the hand and arm | 1 (0.3) | 0 (0) |
| | Systemic | Headache | 26 (7.3) | 26 (7.3) |
| | | Hypertension | 13 (3.7) | 18 (5.1) |
| | | Diarrhea | 7 (2.0) | 11 (3.1) |

| | | | API Batch-1 | API Batch-2 |
|---|---|---|---|---|
| **Total (%)** | | | **354 (100.0)** | **354 (100.0)** |
| | | Cough | 2 (0.6) | 4 (1.1) |
| | | Asthenia | 3 (0.8) | 2 (0.6) |
| | | Nausea | 4 (1.1) | 1 (0.3) |
| | | Upper Acute respiratory infection | 5 (1.4) | 9 (2.5) |
| | | Allergic rhinitis | 1 (0.3) | 4 (1.1) |
| | | Migraine | 3 (0.8) | 0 (0) |
| | | Drowsiness | 2 (0.6) | 0 (0.0) |
| | | Nasal secretion | 1 (0.3) | 3 (0.8) |
| | | Dizziness | 1 (0.3) | 2 (0.6) |
| | | Abdominal pain | 5 (1.4) | 6 (1.7) |
| | | General discomfort | 1 (0.3) | 1 (0.3) |
| | | Vertigo | 1 (0.3) | 2 (0.6) |
| | | Atopic dermatitis | 1 (0.3) | 3 (0.8) |
| | | Sinus bradycardia | 0 (0.0) | 2 (0.6) |
| | | Pharyngeal burning | 0 (0.0) | 1 (0.3) |
| | | Toothache | 2 (0.5) | 0 (0.0) |
| | | Paraesthesia | 1 (0.3) | 0 (0.0) |
| | | Vomiting | 2 (0.6) | 0 (0.0) |
| | | Food Allergy | 1 (0.3) | 1 (0.3) |
| | | Chest pain | 1 (0.3) | 1 (0.3) |
| | | Chills | 1 (0.3) | 1 (0.3) |
| | | Pharyngitis | 1 (0.3) | 1 (0.3) |
| | | Acute ingestion | 1 (0.3) | 1 (0.3) |
| | | Generalized itching | 1 (0.3) | 2 (0.6) |
| | | Blush | 1 (0.3) | 1 (0.3) |
| | | Vagal syndrome | 0 (0.0) | 2 (0.6) |
| | | Anxiety | 1 (0.3) | 0 (0.0) |
| | | Volume increase in the abdominal region | 1 (0.3) | 0 (0.0) |
| | | Insect bite pain | 1 (0.3) | 0 (0.0) |
| | | Edema in lower limbs | 1 (0.3) | 0 (0.0) |
| | | Bilateral epicondylitis | 1 (0.3) | 0 (0.0) |
| | | Multiform erythema | 1 (0.3) | 0 (0.0) |
| | | Glossitis | 1 (0.3) | 0 (0.0) |
| | | Hydroadenitis | 1 (0.3) | 0 (0.0) |
| | | Conjunctivitis | 2 (0.6) | 1 (0.3) |
| | | Acute lower limb lymphangitis | 1 (0.3) | 0 (0.0) |

| | | | API Batch-1 | API Batch-2 |
|---|---|---|---|---|
| **Total (%)** | | | **354 (100.0)** | **354 (100.0)** |
| | | Pain by vesicular lithiasis | 1 (0.3) | 0 (0.0) |
| | | Chest pain | 1 (0.3) | 1 (0.3) |
| | | Eye discomfort | 1 (0.3) | 0 (0.0) |
| | | Lung and pancreas neoplasm | 1 (0.3) | 0 (0.0) |
| | | Upper limb paresthesia | 2 (0.6) | 2 (0.6) |
| | | Dental abscess | 0 (0.0) | 1 (0.3) |
| | | Oral thrush | 0 (0.0) | 1 (0.3) |
| | | Burning when urinating | 0 (0.0) | 1 (0.3) |
| | | Scabies | 0 (0.0) | 1 (0.3) |
| | | Acute atrial fibrillation | 0 (0.0) | 1 (0.3) |
| | | Right knee furuncle | 0 (0.0) | 1 (0.3) |
| | | Bloody stools | 0 (0.0) | 1 (0.3) |
| | | Lipothymia | 0 (0.0) | 1 (0.3) |
| | | Myalgia | 10 (2.8) | 9 (2.5) |
| | | Pneumonia | 1 (0.3) | 1 (0.3) |
| | | Osteochondritis | 0 (0.0) | 1 (0.3) |
| | | Paronychia | 0 (0.0) | 1 (0.3) |
| | | Polaquyuria | 0 (0.0) | 1 (0.3) |
| | | Itching in upper limb | 0 (0.0) | 2 (0.6) |
| | | Salpingitis | 0 (0.0) | 1 (0.3) |
| | | Rectal bleeding | 0 (0.0) | 1 (0.3) |
| | | Tachycardia | 0 (0.0) | 1 (0.3) |
| | | Stiff neck | 0 (0.0) | 1 (0.3) |

Footnote: API: Active Pharmaceutical Ingredient

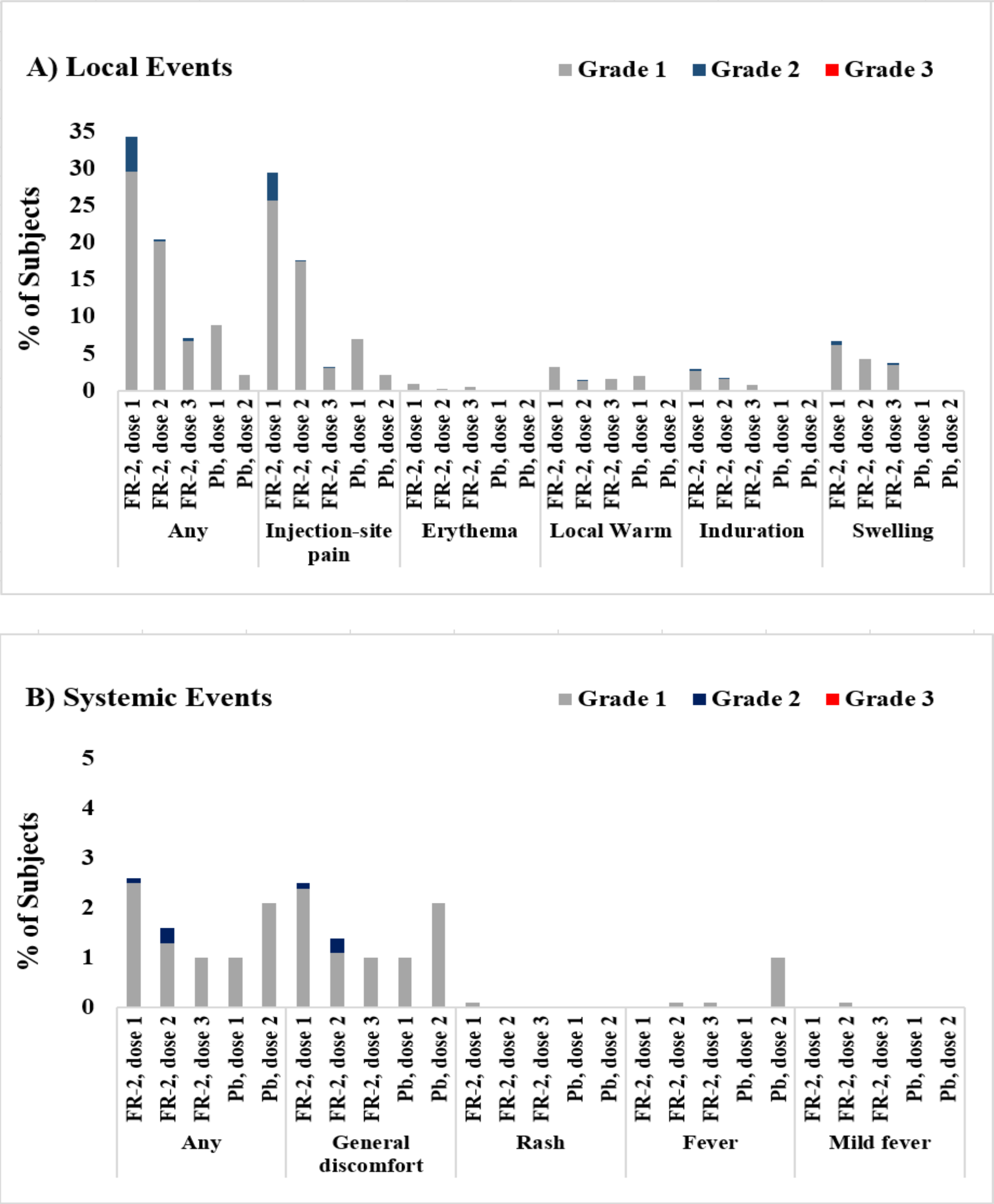


**Figure 1.** Solicited local and systemic events after each dose.

Footnote: Subjects received two doses (on days T0, T28) of SOBERANA 02 (FR-2) or Placebo (Pb) and a third dose (on day T56) of SOBERANA Plus.

**Vaccine** **Placebo**

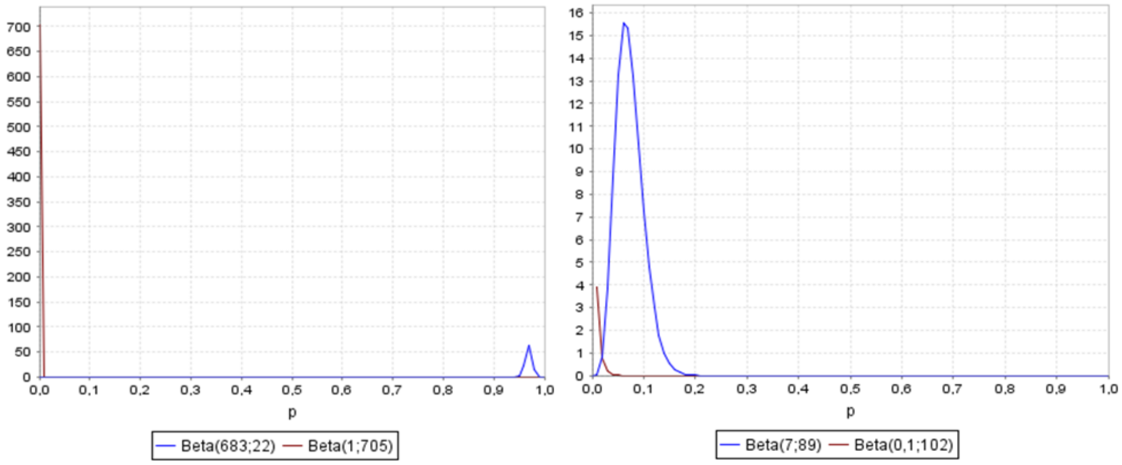


**Bayes Factor (likelihood ratio): 968.0** **Bayes Factor (likelihood ratio): 73.0**

**Figure 2.** Risk-Benefit Balance

Footnote: $Benefit_1$ = Proportion of individuals with seroconversion (≥4-fold increase in antibody concentration over pre-immunization levels) on day 84. $Risk_1$ = Serious vaccine-related adverse events.

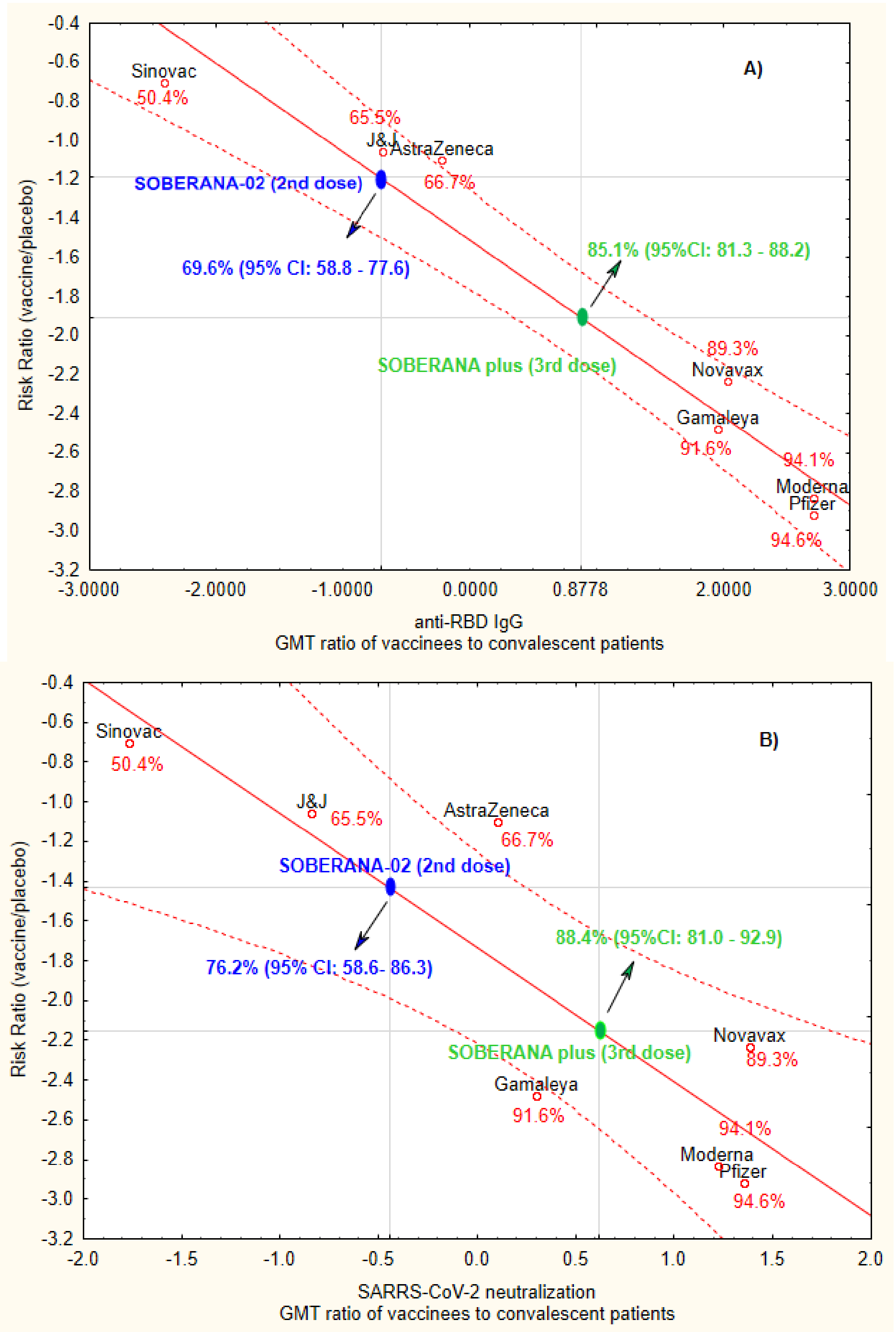


**Figure 3.** Prediction of clinical efficacy from correlation between antibody responses and efficacy rate

Footnote: Panels A and B display correlations of antibody responses for IgG and neutralization and assay ratios, respectively for eight vaccines. The y-axis is estimated log risk ratio reported on the vaccine efficacy scale. The x-axis is log ratio of the peak geometric mean neutralization titer or ELISA titer at 7-28 days post-vaccination, relative to HCS (except for Oxford/AZ and SOBERANA antibody responses, which represent ratios of median titers). Prediction values for SOBERANA were estimated using a regression linear model

### Appendix C. Immunological Techniques

*C.1 Anti-RBD IgG response:* Anti-RBD IgG in sera was evaluated by a quantitative ultramicro ELISA (UMELISA SARS-CoV-2 anti- RBD, Center for Immunoassay, Havana, Cuba) using d-RBD as coating antigen (4 µg/mL) and an in-house standard- characterized serum, which was arbitrarily assigned 200 AU/mL (based on a half-maximal inhibitory titer of 200 and a conventional virus neutralization titer of 160). The standard curve comprised six two-fold serial dilutions (0, 4, 8, 16, 32 and 64 AU/mL) of the standard. Samples were evaluated in duplicate. After incubation step, biotin-conjugate anti-IgG human (0.1 µg/mL) (Sigma Aldrich, San Luis, EE UU) and later, streptavidin/alkaline-phosphatase Roche, Basel, Swiss) in appropriate buffers were added. The final fluorimetric reaction was induced by adding the substrate 4-methylumbelliferyl phosphate (Sigma Aldrich, San Luis, EE UU). The reference curve was constructed using a linear interpolation function. The concentration of anti-RBD IgG was expressed as AU/mL. The seroconversion rate was calculated by dividing the concentration at each time point (at Tx) by the pre-vaccination concentration (at T0). A rate ≥ 4 was considered as seroconversion.

#### *C.2 Molecular viral neutralization test*

This ELISA is an *in-vitro* surrogate of the life-virus neutralization [i]. It uses recombinant RBD-mouse-Fc (RBD-Fcm) and the host cell receptor hACE2-Fc (ACE2-Fch) as coating antigen. Human antibodies against RBD can block the RBD-Fcm interaction with ACE2-Fch. The RBD-Fcm that was not inhibited can bind to ACE2-Fch, and is recognized by a monoclonal antibody anti-γ murine conjugated to alkaline phosphatase. The results are expressed as % inhibition of RBD-hACE2 interaction (at a serum dilution of 1/100); and as the half molecular virus neutralization titre ($mVNT_{50}$) represented as the maximal serum dilution inhibiting 50% of RBD-hACE2 interaction

#### *C.3 Conventional Viral neutralization test*

Neutralizing antibodies against live SARS-CoV-2 was performed in a biosecurity laboratory level 3 (National Civil Defence Research Laboratory, Havana, Cuba) by the conventional virus neutralization test, the gold standard for determining antibody efficacy against SARS-CoV-2, following the recommendation of Manenti & cols [ii]. Serial dilutions of heat-inactivated serum samples (starting from 1:5) in Eagle's Minimal Essential Medium (Gibco, UK) containing 2 % fetal bovine serum (Capricorn, Germany) were incubated for 1 hour at 37°C with an equal volume of viral solution containing 100 $TCID_{50}$ of SARS-CoV-2 (strains: CU2010-2025, variant D614G; CU2101-2102, variant B.1.1.7 alpha; CU2104-2179, variant B.1.617.2 delta; CU2104-2180, variant B.1.351 beta; Cuban Collection at the National Civil Defence Research Laboratory) in cell plates containing a semiconfluent Vero E6 monolayer ($10^4$ cell/well). The highest serum dilution showing an OD at 540 nm, representing the 50% of average OD values from control cell wells (Vero E6 monolayer with mixture of virus-serum) was considered as the neutralization titre and is represented as neutralizing titre 50 ($cVNT_{50}$).

[i] Tan CW, Chia WN, Qin X, et al. A SARS-CoV-2 surrogate virus neutralization test based on antibody-mediated blockage of ACE2–spike protein–protein interaction. Nat Biotechnol. 2020;38:1073-8. https://doi.org/10.1038/s41587-020-0631-z

[ii] Manenti A, Maggetti M, Casa E, et al. Evaluation of SARS-CoV-2 neutralizing antibodies using a CPE-based colorimetric live virus micro-neutralization assay in human serum samples. J Med Virol. 2020; 92(10): 2096-104. https://doi.org/10.1002/jmv.25986
